# Linked Exposures Across Databases (LEAD): An exposure data aggregation framework to facilitate clinical exposure review

**DOI:** 10.1101/2024.03.01.24303567

**Authors:** Immanuel B.H. Samuel, Kamila Pollin, Sherri Tschida, Calvin Lu, Michelle Prisco, Robert Forsten, Jose Ortiz, John Barrett, Matthew Reinhard, Michelle Costanzo

## Abstract

Understanding the health outcomes of military exposures is a critical effort for Veterans, their health care team, and national leaders. Veterans Affairs providers receive reports of military exposure related concerns from 43% of Veterans. Understanding the causal influences of environmental exposures on health is a complex task advancement in exposure science and may require interpreting multiple data sources; particularly when exposure pathways and multi-exposure interactions are ill-defined, as is the case for complex and emerging military service-related exposures. Thus, there is a need to standardize clinically meaningful exposure metrics from different data sources to guide clinicians and researchers with a consistent model for investigating and communicating exposure risk profiles. The Linked Exposures Across Databases (LEAD) framework provides a unifying model for characterizing exposure from different exposure datatypes and databases with a focus on providing clinically relevant exposure metrics. Application of LEAD is demonstrated through comparison of different military exposure data sources: Veteran Military Occupational and Environmental Exposure Assessment Tool (VMOAT), Individual Longitudinal Exposure Record (ILER) database and a military incident report database, the Explosive Ordnance Disposal Information Management System (EODIMS). This cohesive method for evaluating military exposures leverages established information with new sources of data and has the potential to influence how military exposure data is integrated into exposure health care and investigational models.

## Introduction

Understanding the health effects of military exposures is a top priority for clinicians and researchers focused on Veteran health care, with 43% of Veterans reporting toxic exposure concerns to their Veterans Affairs (VA) health care providers (TES, n.d.). The recent passing of The Sergeant First Class Health Robinson Honoring our Promise to Address Comprehensive Toxics Act (PACT Act) in 2022, expands VA health care and benefits, and expands presumptive health conditions resulting from various military combat area deployments and exposures. This significant increase in military toxic exposure interest requires integration of appropriate data to support investigations of health outcomes of military exposures. Capturing the exposome, which assesses the multifaceted associations between environment, behavior, biology, and disease over time is necessary (Chung et al., 2021) as exposures do not end after the military. Utilizing the exposome model to better research military toxic exposures is crucial as it considers the totality of environmental influences on individuals across the lifespan (Stingone et al., 2017). By examining not only traditional combat-related exposures but also broader factors that influence the exposome such as psychosocial stress and other exposures, the exposome provides a comprehensive understanding of complex interactions shaping health outcomes in military personnel. The approach presented in the LEAD framework can support efforts to understand the exposome.

To support the clinical and research surveillance and investigational efforts, many tools, programs and data sources have been developed. These efforts include the VA Toxic Exposure Screen (TES, n.d.) which is a brief system-wide VA questionnaire that broadly assesses Veterans’ military related exposure concerns and is conducted at multiple time points. Additional individual exposure data sources exist in the VA and Department of Defense (DoD), such as VA registry exams (Agent Orange Registry, Airborne Hazards and Open Burn Pit Registry, Gulf War Registry, Ionizing Radiation Registry, Depleted Uranium Follow-Up Program, Toxic Embedded Fragment Surveillance Center), Post-Deployment Health Assessment (PDHA), Re-assessment (PDHRA) (Griffith, 2022), and Defense Occupational and Environmental Health Readiness System (DOEHRS) (Krahl et al., 2022; DHA), n.d.). To integrate information into a single source, the DoD and VA have developed the Individual Longitudinal Exposure Records (ILER) (Bradburne & Lewis, 2017; ILER, n.d.) for each service member and Veteran which includes information beginning with entry into the military and spanning across the military career. However, exposure data aggregation by itself without a framework to interpret that information neglects the need to generate clinically insightful exposure profiles and dismisses the utilization of a consensus in exposure data research. Failure to consider the clinical relevance of exposure data may miss an important opportunity to advance exposure-health knowledge (Gochfeld, 2005; Levine, 2006; Nielsen & Ovrebø, 2008). Another benefit for summarizing exposure data sources for clinical purposes is to assist the VA with presumptive conditions. There are certain conditions considered presumptive conditions in which the VA automatically presumes that the condition or disability was caused by military service. For instance, Hodgkin’s disease is a presumptive condition for Veterans exposed to Agent Orange during specific deployments (Health, n.d.). Thus, understanding exposures and health outcomes can aid the VA in their efforts to better comprehend presumptive conditions.

The main challenge in summarizing exposure data is the lack of a broadly defined, clinically relevant, standardized methodology to accommodate sparse information across exposure databases. A framework for collating exposure data is needed to estimate ubiquitous exposure dose metrics that could be used to align with population-level clinical analyses (e.g., mortality and morbidity), research cohort selection (e.g., high exposure vs. low exposure groups), or clinical consultations (e.g., risk communication). Clinicians and researchers often use varying methods to evaluate exposure data, and some may not follow best practices (Velentgas et al., 2013). For this reason, comparisons of exposure assessments from one clinician to another, as well as studies that use nuanced methods of exposure characterization, is increasingly challenging in all settings. For exposure data to be clinically relevant it needs to be evaluated, documented, and summarized based on a standardized model of understanding.

A common framework for information capture and data integration will guide the development of consistent exposure summaries from pooled information from multiple databases. Such an exposure capture framework will support clinicians and researchers to acquire and interpret information more accurately, lead to improved application of exposure knowledge to identify knowledge gaps in exposure characterization, and enhance clinical care. This unified framework for evaluating and interpretating exposure data can serve as a consensus helping establish standardized methods ensuring consistency in research and improving clinical care.

To address this need, the D.C. WRIISC (War Related Illness and Injury Study Center) developed the Linked Exposures Across Databases (LEAD) framework to (1) bridge multiple exposure data sources by defining common data elements to collate information that can then be combined across new and existing assessment tools and databases, and (2) provide a foundation for further summarization of these findings for exposure dose estimation, clinical exposure review and communicate concise exposure history to Veterans.

## Linked Exposures Across Databases (LEAD) framework

The LEAD conceptual framework is designed to accommodate exposure information from a wide range of different sources by defining common exposure data elements that are frequently used across data sources. The LEAD framework was designed to specifically address gaps in exposure information sourcing and exposure characterization with a focus on the clinical utility of exposure data. One key focus area is to enable development of health application for siloed data sources, such as military records and incident reports that are not typically used for clinical evaluations but contain pertinent exposure information. The LEAD framework derives from a commonly used form of dosage calculation which considers total dosage as a function of intensity of exposure, and the time over which that exposure is sustained. This method of calculating exposure using the form of dosage calculation is used across fields such as radiation exposure capture (Lai & Levitt, 2022), toxicity research (Di Credico et al., 2020) and the military to calculate blast overpressure as the product of pressure over a set period (Taylor & Ford, 2009; Ouellet & Philippens, 2017). These applications are focused on specific exposures and are limited in scope to specific research questions and do not provide a comprehensive exposure profile. LEAD’s approach involves broad definitions of common exposure data elements to enable unified exposure characterization across varied exposures while utilizing existing exposure capture paradigms and shaping new research approaches. This enables LEAD to integrate multiple data sources, translate individual and population level exposure data for clinical use and generate toxic exposure surveillance and research data applications.

While quantitative measures of exposure intensity (e.g., Pascals for blast pressure, micrograms for chemical exposure) and time of exposure (measured in seconds) using sensors are the most objective and quantifiable form of measurement, such information is not typically available at the individual level for Veterans concerned about potentially toxic military exposures. Therefore, the LEAD framework aims to expand the application of intensity and time-based dosage estimation to large sets of qualitative and subjective data. Additionally, potential moderating factors such as protective controls are also considered since these factors contribute to exposure-related outcomes.

LEAD characterizes exposure using the following common exposure data elements:

*Exposure Dose f* (*Intensity × Time* | *Moderators*)

*Intensity f* (Route, Proximity, Symptoms)

*Time f* (Duration, Frequency, Period)

*Moderators f* (Environmental Controls, Personal Protective Controls)

Given the sparse availability of objective exposure information, the LEAD framework can enable aggregate exposure dosage estimation using proxy measures of exposure intensity and time, as well as estimates of factors that may moderate these effects. The LEAD framework reflects our aim to characterize exposure consistently. However, just as in current clinical practice where some exposure features are more important given their perceived effects on outcomes, the exposure variables above must be assigned weights accordingly when assessing exposure dose. In this report we provide exposure expert informed weights that reflect the importance given to specific variables while evaluating exposures in the VA for the purpose of demonstrating a practical example of exposure dose estimation. Future analysis on using LEAD exposure data with health outcomes data would enable empirical weight assignment using risk-modeling approaches (e.g. Cox-Proportion Hazards Model).

**Figure 1:**
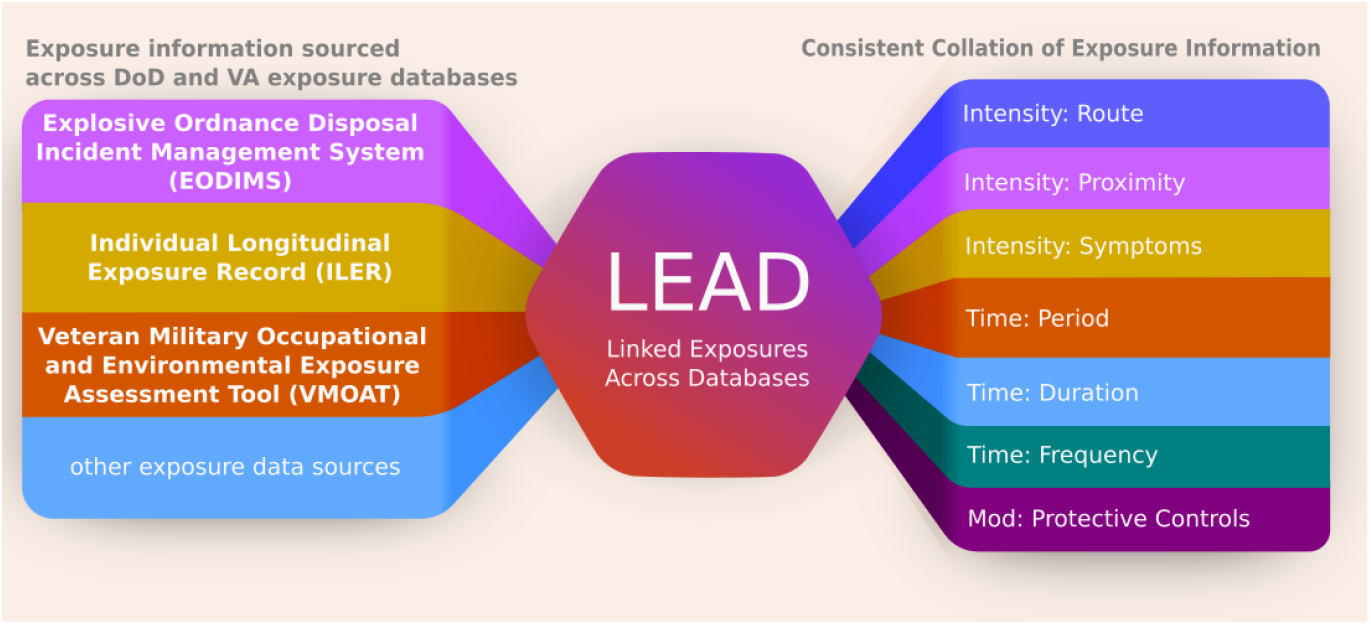
LEAD framework is designed to accommodate exposure information from a wide range of different information sources by defining common exposure data elements to obtain a weighed exposure metric for further analysis and clinical use.

To illustrate how the LEAD framework can be used to consolidate exposure data into a unifiable metric, this report will present information from three different exposure data sources relevant to a cohort of Explosive Ordnance Disposal (EOD) Veterans, a complex and very high environmental exposure military occupation. We provide sample data extraction of exposure information from multiple databases and their collation in Table 1. Further, example of how the data can be used to compare high and low blast exposure is provided in Table 2, and one possible method of scoring such information is provided in Table 3.

**Table 1:** LEAD exposure collation overview: A sample extraction of exposure data available across multiple exposure databases. Exposures reported in this table are not tied to an individual but are pulled from representative sources to provide an example of how the LEAD framework can be used to collate information.

| 1. Exposure Collation |  | Intensity |  |  | Time |  |  | Moderators |
| --- | --- | --- | --- | --- | --- | --- | --- | --- |
| Source | Exposure | Route | Proximity | Symptoms | Period | Duration | Frequency | Protective Controls |
| EODIMS | Blast | Impact | 10 feet | Lost consciousness | 03/01/1999 – 03/02/1999 | 4 hours | 1 event | Suit |
| EODIMS | Blast/50lbs NEW | Impact | >150m | None | 07/01/2000 – 07/02/2000 | < 2 minutes | 1 event | Bunker |
| EODIMS | Blast/<5lb NEW | Impact | < 15 m | n/a | 11/01/2000 – 11/02/2000 | 4 hours | 1 event | Safe area |
| VMOAT | Blast | Impact | 10 feet/ | Lost consciousness | 03/01/1999 – 03/02/1999 | 4 hours | Frequent | Suit |
| ILER-PDHA | Vehicle Accident | Impact | n/a | Neck/back injury; LOC <5 mins; Disorientation | 02/11/2000 – 02/11/2000 | n/a | 1 event | Seat belt |
| ILER-PDHA | Doxycycline (Vibramycin®) | Ingestion | n/a | n/a | 02/01/2000 – 11/01/2000 | 6 months | daily | n/a |
| ILER-PDHR-A | Sand/Dust | Inhalation | n/a | Coughing, difficulty breathing | 04/01/2000 – 01/01/2001 | 10 months | n/a | n/a |
| ILER-EIR | Fuel | Skin, Inhalation | 10 feet | Rash | 03/01/1999 – 03/02/1999 | 3 hours | Less Frequent | Mask, Ventilation |
| ILER-POEMS | Food/water borne disease | Ingestion | n/a | High risk: bacterial diarrhea, hepatitis A, typhoid fever<br>Moderate risk: diarrhea-cholera, diarrhea-protozoal, brucellosis and hepatitis E | 04/01/2000 – 01/01/2001 | daily | less frequent | Hepatitis A and typhoid vaccination<br>Food/water consumption only from approved sources |

**Table 2:** Sample high / low exposure data extracted using the LEAD framework shows available data for a specific exposure type (blast) for two mock EOD Veterans. Exposures reported are not tied to an individual but pulled from representative sources to provide an example of how the LEAD framework can be used to compare differences in exposure dose for a specific exposure.

| 2. Blast Exposure Collation |  | Intensity |  |  | Time |  |  | Moderators |
| --- | --- | --- | --- | --- | --- | --- | --- | --- |
| Source | Exposure | Route | Proximity | Symptoms | Period | Duration | Frequency | Protective Controls |
| Low Exposed |  |  |  |  |  |  |  |  |
| EODIMS | Blast | Skin Contact | 250 feet | None | 03/01/1999 – 03/02/1999 | 4 hours | 1 event | Suit |
| VMOAT | Blast | Skin Contact (dust) | within 500 feet | None | 05/01/1985 – 03/02/1999 | 4 hours | Infrequent (once a year or less) | Gloves, Eyewear, Headgear |
| ILER-PDHA | Blast or Explosion (IED, RPG, EFP, land mine, grenade, etc.) | Impact | More than 100 meters (328 feet) | None | 03/01/1999 – 03/02/1999 | n/a | 1 event | n/a |
| High Exposed |  |  |  |  |  |  |  |  |
| EODIMS | Blast | Impact, Inhalation | 25 feet | Lost consciousness | 03/01/1999 – 03/02/1999 | 4 hours | 1 event | Suit |
| VMOAT | Blast | Impact, Inhalation, Skin Contact, Eye Contact | within 50 feet | Severe (Lost consciousness 30minutes-24 hours) | 01/01/1984 – 10/01/1994 | 2-4 hours per day | Frequent (Daily) | None |
| ILER-PDHA | Blast or Explosion (IED, RPG, EFP, land mine, grenade, etc.) | Impact (not directly listed) | Less than 25 meters (82 feet) | LOC 5-30 min, Seeing stars | 03/01/1999 – 03/02/1999 | n/a | more than 3 (5 times) | n/a |

**Table 3:** Lookup table used to translate exposure information in table 2 to categorical scores as shown in table 4.

| 3. Scoring | Intensity |  |  | Time |  |  | Moderators |
| --- | --- | --- | --- | --- | --- | --- | --- |
|  | Route | Proximity | Symptoms | Period | Duration | Frequency | Protective Controls |
| 0 | na | na | none | na | na | na | none |
| 1 | 1 route | >250m | 1 | 1 day - 1 week | <1 hour | 1 event | 1 |
| 2 | 2 routes | 100m - 250m | 2 | 1 week - 1 month | 1-2 hours | 2 events | 2 |
| 3 | 3 routes | 10m - 100m | 3 | 1 month - 1 year | 2-4 hours | 3 events | 3 |
| 4 | 4 routes | 1m - 10m | 4 | 1 year - 10 years | 4-8 hours | >4 events | 4 |
| 5 | >5 routes | <1m | 5 | >10 years | >8 hours | routine exposure | >5 |

Data sources include:

1. **Explosive Ordnance Disposal Information Management System (EODIMS):** The EODIMS System is an operations specific Classified and Unclassified program of record incident reporting system with discoverable data database managed by the Air Force supporting the joint service EOD units. Focused use of these records for clinically relevant exposure data enables access to the immediate post-exposure reports that are less affected by recall bias and may represent direct evidence of a possible hazardous exposure.
2. **Veteran Military Occupational and Environmental Exposure Assessment Tool (VMOAT):** The VMOAT is a self-reported, structured, and lifespan-based comprehensive assessment of occupational exposures sustained during military service as well as during non-uniformed military and civilian work periods. The VMOAT includes a demographic information section, a lifespan-based occupational and environmental history section, and a comprehensive exposures section based on evidence-based exposure categories such as chemical, physical, injuries, biological, and psychological (Samuel et al., 2022).
3. **Individual Longitudinal Exposure Record (ILER):** The ILER is an individual, electronic record of exposures for each service member and Veteran. ILER contains information from other sources including deployment dates and locations, all-hazard occupation data, environmental hazards, objective monitoring, medical encounter information and medical concerns regarding possible exposures. ILER aims to deliver capabilities and improvements in health care, benefits, collaborations between VA, DoD, Congress, beneficiaries, and other stakeholders (such as Veterans Service Organizations), as well as research and integration of exposure data from VA’s environmental health registries (ILER, n.d.).

## Application of the LEAD to collate exposure variables across exposure databases

Integrating data into exposure variables to be analyzed when determining exposure dose across various hazard categories is a primary function of LEAD. Exposures are categorized according to domains: chemical, biological, physical, ergonomic/injury, and psychological hazards, as defined by the Department of Labor’s Occupational Safety and Health Administration (OSHA, n.d.). A description of each of these variables is detailed, along with examples of exposure information available across EODIMS, VMOAT and ILER.

### 1. Intensity

Exposure intensity can be directly estimated by assessing the amount or concentration of hazardous substances that an individual comes into contact. In most cases, intensity exposure characterization occurs retrospectively with limited quantitative exposure data. In such cases, indirect measures of exposure or proxies for intensity such as routes of exposure, proximity to exposure source, and symptoms at the time of the exposure event are the only means of assessing intensity. Such indirect estimates supplement objective information that is often not available.

#### 1.1 Route

The route of exposure refers to how a substance enters the body. The EODIMS database does not contain extensive documentation of exposures routes, but routes may be inferred from the incident type and specific exposures reported. For example, an incident report documenting post-blast aerosolized particulate matter may be inferred to have inhalation and skin contact as possible exposure routes. The VMOAT categorizes exposure routes into the following categories: inhalation, ingestion, skin/eye contact, injection. Psychological exposures routes are categorized as experiencing, seeing, and/or hearing. ILER contains location and group-level exposure records that may be associated with chemical (e.g., burn pits) and biological (e.g., infections) exposures. While group level estimates cannot be directly interpreted at the individual level, these estimates will be used as a probabilistic estimate within LEAD.

#### 1.2 Proximity

Proximity to the exposure source is associated with exposure intensity since typically the closer an individual is to the hazard, the greater the exposure intensity, leading to a higher dose. Report data captured in EODIMS may offer proximity information including distances between safe areas and incident sites and frequency of travel between these areas, and other locations involved in the documented response. While the initial VMOAT did not estimate proximity, the VMOAT 2.0 will assesses proximity to exposures. Estimates of proximity to exposures can also be gathered from ILER in sources such as VA registries, but these are limited in scope and depth of information collected such as asking binary yes/no questions (e.g., whether the individual was near a burn pit, sewage pit, blast from an IED or other exposures).

#### 1.3 Symptoms at the time of exposure

Presence of medical signs and symptoms following exposure suggests higher exposure intensities, for many exposures. While the purpose of LEAD is not to assess health outcomes, assessing the presence of health changes immediately following exposure can be used as a proxy for estimating exposure intensity especially when objective exposure intensity data is unavailable. We note here that the absence of symptoms or the lack of documentation of symptoms should not be interpreted as a lack of exposure, as devastating health consequences may occur years after exposures (e.g., mesothelioma in asbestos workers) (Selikoff et al., 1980; Seidman et al., 1982; for Disease Control & CDC, n.d.). While EODIMS documents capture immediate health effects associated with each exposure incident, such capture is neither consistent nor uniformly documented. While ILER contains some records that ask deployment health effects questions (eg. PDHRA accessed through ILER asks about concussions during deployment) and additional health outcomes data may be included in the future in a limited capacity.

### 2. Time

The timing of exposure is important and can be directly estimated with a variety of approaches, ranging from sensors with high sampling rates to subjective reports. Data at the individual level though is sparse and often requires interpreting subjective narratives of exposures that are incomplete and can vary from person to person (for example, some individuals can recall in detail the timing of an exposure whereas others will report a general time during a deployment). Duration and frequency of an exposure can be used to estimate exposure timing.

#### 2.1 Duration and Frequency

The duration of exposure refers to how long someone is exposed to a substance or hazard. The longer the duration, such as noise (Kaufman et al., 2005), or long-term bio accumulation of Polyfluoroalkyl substances (Seals et al., 2011; Ankley et al., 2020), generally the greater the total dose and risk of adverse outcomes. Similarly, frequent repeated exposures even at smaller intensities, such as repeated blast exposures, can lead to chronic health effects (Wang et al., 2020). Duration and frequency are closely related, yet how these variables are assessed is inconsistent across databases since there are no well-established standards for such documentation. The EODIMS database contains detailed time and frequency estimates such as start and completion times whereas the VMOAT assesses ‘duration’ in the context of the number of hours during a day and ‘frequency’ is assessed in events within a time period. ILER does not contain distinct duration and frequency estimates in most cases, but some exposure Registry assessments assess hours in a typical day or days in a typical month an individual may have been exposed (e.g., airborne hazards including burn pits, fumes, dust or other similar exposures).

#### 2.2 Period or the Time of Exposure

The exposure period is associated with occupational history or military deployments. The start and end dates for occupational periods are assessed either by asking for the start and end times for exposures in questionnaires, as done in VMOAT, or through administrative records contained in the ILER or EODIMS. Since this variable records the time of exposure, combined with date of birth, this variable can be used to estimate (i) age at exposure, (ii) time since exposure and (iii) cohort effects across military eras which are key factors that may affect outcomes.

### 3. Moderators

Hazard controls play an important role in moderating occupational exposure (Hymel et al., 2011). While individual factors that affect exposure tolerance (Larson et al., 2008), are important moderating factors, they are not assessed broadly. Given the growing body of literature on the exposome and potential health outcomes (Siroux et al., 2016; Vermeulen et al., 2020) it is important that exposure assessments incorporate these contributing factors as exposure science develops.

#### 3.1. Environmental and Personal Protective Controls

The National Institute of Occupational Safety and Health (NIOSH) hierarchy of controls (NIOSH, n.d.) has been developed to control worker exposures, reduce or remove hazards. and reduce risk of illness or injury. When elimination or substitution (I.e. most effective on the hierarchy) of the hazard is not possible, engineering controls such as using ventilation systems administrative controls such as implementation of rotating work schedules and Personal Protective Equipment (PPE) can reduce exposures (Reddy et al., 2019). The EODIMS contains detailed logs of the individual use of such hazard controls in each incident report, as well as other workplace controls. While detailed assessment of exposure control measures is not practical due to the retrospective and subjective nature of the VMOAT, the survey does assess the use of PPE at the individual level. ILER does document Hazard Controls in DOEHRS Industrial Hygiene (IH) reports, but these reports are typically cohort level reports.

### 4. Examples of How to Use LEAD framework

Drawing from the above components of the LEAD framework, Tablets 1-4 illustrate the process of collating exposure information across multiple sources into a consistent format (Table 1) and provides a preliminary scoring method (Table 3 and 4) for blast exposure data (simulated) based on a typical EOD Veteran exposure profile (Table 2).

**Table 4:** Dose Estimation: A sample scoring of blast exposure data across multiple exposure databases for two mock EOD Veterans. The scoring is based on weights assigned by exposure experts meant to represent the clinical significance of the exposure data that has been numerically represented from Table 2. Note: data on individual differences is sparse at present, so this example is representative of the types of information that are currently available. Future efforts to estimate individual differences data from other health assessment systems is needed since that is an important aspect of exposure dose estimation.

| 4. Dose Estimation | Variables |  |  |  |  |  |  | Weighted Score |
| --- | --- | --- | --- | --- | --- | --- | --- | --- |
|  | Route | Proximity | Symptoms | Period | Duration | Frequency | Protective Controls |  |
| Expert Informed Weights | 1 | 1 | 5 | 1 | 2 | 2 | -2 |  |
| Blast Exposure Data |  |  |  |  |  |  |  |  |
| Low Exposed Example |  |  |  |  |  |  |  |  |
| EODIMS | 1 | 1 | 0 | 1 | 3 | 1 | 1 | 9 |
| VMOAT | 1 | 1 | 0 | 4 | 3 | 1 | 3 | 8 |
| ILER-PDHA | 1 | 2 | 0 | 1 |  | 1 |  | 6 |
|  |  |  |  |  |  |  | Average: | 7.67 |
| High Exposed Example |  |  |  |  |  |  |  |  |
| EODIMS | 2 | 4 | 3 | 1 | 3 | 1 | 1 | 28 |
| VMOAT | 4 | 3 | 4 | 4 | 3 | 5 | 0 | 47 |
| ILER-PDHA | 1 | 4 | 2 | 1 |  | 4 |  | 24 |
|  |  |  |  |  |  |  | Average: | 33.00 |

## Discussion

The LEAD framework offers a means to create an aggregate of exposure information across diverse military and non-military sources which can then be synthesized to generate meaningful exposure metrics for clinical use. The value of this evidence-based approach has direct clinical and research relevance to optimize how exposure risk factors may influence the development of disease.

Current exposure assessment tools are limited in scope and require review of data from varied assessments and need high level of exposure literacy. Not all VA care providers can be expected to have comprehensive knowledge of exposure assessment methodologies and interpret available data in a consistent way. The LEAD provides a consistent framework and enables simplified exposure metrics which can be used for quick clinical review without extensive exposure knowledge. This does not invalidate existing nuanced exposure assessment methodologies which may be required in specialized exposure assessment clinics. The common data elements-based exposure evaluation model facilitates consistency in exposure communication between care providers as well as consistency in interpretation of exposure data. The preliminary estimation of simplified exposure metrics presented in this report along with future data-driven empirical dose estimates will further enable consistency in exposure evaluation and could reduce variability in interpretation of exposure data. The goal for LEAD is to improve exposure profiling and provide a practical template to collate information across different exposure sources to drive meaningful insights for clinicians and researchers.

Limitations: While each of these three databases are valuable in providing exposure information as observed in Table 1, there are limitations to these databases which emphasizes the need for collating information across each of them. EODIMS provides documented accounts of occupational incidents that involves health risks, and contains significant data such as occupation, deployment data, exposure descriptions, and biometric data. Since, much of the data in EODIMS is classified, extensive work is required to redact the data for healthcare related use in the VA. The LEAD framework provides a systematic method for targeted extraction of non-operational health-relevant information for exposure assessment and care at VA clinics. Importantly, while the current application of LEAD has been to integrate EODIMS data to create exposures profiles for EOD Veterans, it is designed to generalize exposures from other military occupations as well as civilian exposures.

The VMOAT is a subjective survey and, while it is comprehensive in nature, takes approximately 45 minutes to complete. VMOAT is not a screening tool as it is designed to be administered to Veterans with complex exposure histories and thus it is not practical to administer to all Service Members and Veterans. While VMOAT provides important self-reported exposure information, the challenge lies in integrating this information with other exposure records with varying information into a consistent and easily understandable format. LEAD aims to provide a path for integration of VMOAT data with other exposure databases.

ILER aims to integrate individual and population level exposure data from multiple existing operational databases within the VA and DoD. However, ILER data is often presented in a format understandable by occupational medicine providers and industrial hygienists only since ILER contains jargon that makes it difficult to understand. ILER information is not prioritized which makes clinically relevant information extraction tedious. Information at the population level may not correlate with the level of exposure an individual service member had encountered. For this reason, information from other sources, such as EODIMS and VMOAT, are required to elicit potential population level exposures and LEAD allows merging across these sources for a wholistic interpretation of exposure data.

Another limitation is that while an expert informed weighted scoring methodology is outlined in Tables 2 and 3, empirical weight assignment using health outcome metrics is required to calculate evidence-based estimation of exposure scores. Use of such methods will allow for outcome specific weight estimation leading to outcome specific exposure risk scores, and advance exposure pathways science. Additionally, individual differences in genetics and existing health conditions at the time of exposure influence exposure tolerance. However, such data is typically not assessed along with exposure assessments. Future iterations of this framework aim to include the VA and DoD health records to provide data driven weight estimation and identify other potential moderating factors such as health status at the time of exposure.

## Conclusion

To address Veteran exposure concerns, the VA should improve models aimed at improving our knowledge of military occupational exposures and its potential impact on health. This can be accomplished by collaborating with the DoD and other partners to ensure military exposure data is formatted to in a way that supports Veteran clinical exposure assessments. Through subject matter expertise and review of the occupational and environmental medicine literature, the LEAD framework defines common exposure data elements for collating exposure information and provides standardized elements for extraction from disparate and siloed DoD and VA databases. These exposure data elements are essential building blocks for creating consistent, succinct, insightful, comprehensive, and clinically relevant exposure profiles. Incorporating the common exposure data elements outlined in this framework in future exposure studies enables consistency, comparability, and robustness in data collection and analysis across exposure studies. This framework will also avoid inappropriate or improper interpretations of exposure data when evaluating Veteran exposures.

LEAD aligns with the VA’s PACT ACT directives by offering a path forward to understand how hazardous exposures may impact Veteran health and assist the VA to identify new presumptive conditions for Veteran care and benefits. The long-term goal for the LEAD framework is to create consistent and clinically relevant exposure summaries from information across multiple data sources to optimize clinical and research processes associated with exposure data acquisition and use.

## Data Availability

This report features an investigational framework for reviewing exposures for clinical use using simulated data. Data were simulated due to the sensitive nature of the underlying information.

